# Genetic risk factors associated with SARS-CoV-2 susceptibility in multiethnic populations

**DOI:** 10.1101/2022.06.23.22276797

**Authors:** Aditya Dandapani Sriram

## Abstract

Susceptibility to infection from severe acute respiratory syndrome coronavirus 2 (SARS-CoV-2), the virus that causes the disease COVID-19, may be understood more clearly by looking at genetic variants and their associations to susceptibility phenotype. I conducted a genome-wide association study of SARS-CoV-2 susceptibility in a multiethnic set of three populations (European, African, and South Asian) from a UK BioBank clinical and genomic dataset. I estimated associations between susceptibility phenotype and genotyped or imputed SNPs, adjusting for age at enrollment, sex, and the ten top principal components of ancestry. Three genome-wide significant loci and their top associated SNPs were discovered in the European ancestry population: *SLC6A20* in the chr3p21.31 locus (rs73062389-A; P = 2.315 × 10-12), *ABO* on chromosome 9 (rs9411378-A; P = 2.436 × 10-11) and *LZTFL1* on chromosome 3 (rs73062394; P = 4.4 × 10-11); these SNPs were not found to be significant in the African and South Asian populations. A multiethnic GWAS may help elucidate further insights into SARS-CoV-2 susceptibility.

## Background

Severe acute respiratory syndrome coronavirus 2 (SARS-CoV-2) is a member of the coronavirus family of viruses and specifically causes the disease COVID-19. Individuals infected with SARS-CoV-2 experience a greatly heterogenous range of health outcomes ranging from asymptomatic infection to flu-like symptoms, shortness of breath, loss of smell or taste, and in severe cases, death.^1^ With COVID-19 having such variable symptom presentation and susceptibility patterns, it is becoming increasingly important to understand the mechanisms of infection, divergent symptoms, and how infection response may differ on an individual-to-individual basis. Genetic characterization of SARS-CoV-2 has already provided insights into the virus’s mechanism of infectivity and, in combination with analysis of human host genetic variants, can inform clinicians and geneticists in developing treatment plans to ease the burden of COVID-19.^2^ I conducted a multiethnic genome-wide association study (GWAS) to investigate possible associations between single-nucleotide polymorphisms (SNPs) and susceptibility to SARS-CoV-2 infection.

Several research organizations and initiatives have been at the forefront of tackling the COVID-19 pandemic from a genetics perspective. 23andMe, AncestryDNA, and the National Institutes of Health have released GWAS results for a number of COVID-19 phenotypes of interest.^3, 4, 5^ While more extensive research has been conducted on the severity of SARS-CoV-2 infection, susceptibility is still a relatively unknown and unconfirmed phenotype. The COVID-19 Host Genetics Initiative has consolidated results from three genome-wide meta-analyses consisting of nearly 50,000 patients from 19 countries, reporting 13 loci associated with SARS-CoV-2 infection or COVID-19 phenotype that reached genome-wide significance.^6^ The UK Biobank continues to upload COVID-19 test results to their repository of patient data and updates their online GWAS results portal with any available genetic association results from COVID-19 genetic scans.^7^ Existing GWAS for SARS-CoV-2 susceptibility have shown the chr3p21.31 locus to be strongly associated with both severity and susceptibility phenotypes. The rs2271616:G>T variant has been associated with susceptibility phenotype across multiple studies.^5, 8, 9^

A general concern about genetic association studies is the predominantly European ancestral makeup of study participants. A majority of GWAS across all traits and diseases have been conducted in European-ancestry populations, and it is very important to understand health outcomes and genetics across a broader set of ancestries. By using a diverse set of study populations, this multiethnic GWAS aims to provide updated information regarding potential causal variants for COVID-19 susceptibility across a greater range of individuals.

## Methods

### Data and Sample Information

All data processing and analyses used version 3 of the UK BioBank imputed dataset, consisting of genomic data for 487320 participants of several ancestry groups including 459250 individuals of European ancestry (EUR), 7644 individuals of African ancestry (AFR), 9417 individuals of South Asian ancestry (SAS), and 11009 individuals of other ancestries besides these (OTHERS).^10, 11^ Participant age at enrollment ranged from 37 to 73 years.

The data analyzed consist of three populations with COVID-19 test results from the UK BioBank dataset: EUR (16551 positive test results, 81826 negative test results), AFR (557 positive test results, 1281 negative test results), and SAS (810 positive test results, 1516 negative test results). As per the definition of the UKB dataset criteria for susceptibility, cases were individuals with a laboratory-reported positive test for SARS-CoV-2, and controls were participants who received lab-reported negative test results (labeled “population”).^5^ I looked at SARS-CoV-2 susceptibility as the phenotype of interest for this association study. COVID-19 hospitalization, severity, and death were the three other phenotypes in the dataset that merited further exploration in separate studies. An important point to note is that there is no information as to whether individuals were exposed to SARS-CoV-2. Therefore, results from this association study are primarily concerning information on genetic factors for positive test results. As more participant data are collected, the UK BioBank continues to update the dataset with information about test results and participant COVID-19 phenotypes. This is a cross-sectional rather than case-control study, and COVID-19 test results of the participants are as of June 18th, 2021.

### Analysis - Genotyping, Imputation, GWAS Setup

All UKB individuals were genotyped using the Applied Biosystems(tm) UK Biobank Axiom(tm) Array, consisting of 825927 genetic markers. Imputed genotypes were included using the Haplotype Reference Consortium (HRC) reference panel and 1000 Genomes Project phase 3.^12^ Imputation for this UKB dataset increased the number of markers available for association testing, and improved the statistical power of the GWAS conducted using genetic information from this dataset. The GWAS for each ancestral population was conducted using SAIGE, a Scalable and Accurate Implementation of a Generalized mixed model v0.38.^13^ SAIGE methodology accounts for population stratification, sample relatedness and existing case-control imbalance. In the African and South Asian population GWAS there were very low case numbers compared to the European analysis, so SAIGE was a preferred tool to control for the issue of smaller sample sizes. SAIGE applies a saddlepoint approximation to control for inflation that may arise from unbalanced case-control ratios.^14^ The analysis in each GWAS adjusted for age at enrollment, sex, and the ten top principal components of ancestry. Logistic regression was the implemented regression analysis method.

For the data used in each GWAS, SNPs were filtered based on their imputation quality (in this case, r^2^ > 0.7) and their minor allele frequencies (MAF > 0.01). After filtering and running the SAIGE logistic regression analysis on the data, the calculated lambda (genomic inflation factor) values ranged from 1.022 to 1.055 in the analyses of each separate ancestry group, indicating no concerns about systematic genomic inflation. Quantile-quantile (QQ) plots for each group, shown in Figures 1, 2, and 3, validated the conclusion that there were no discernable signs of genomic inflation. SNPs were considered genome-wide significant if they met the widely-accepted threshold of P < 5 × 10^−8^.

**Figure 1.**
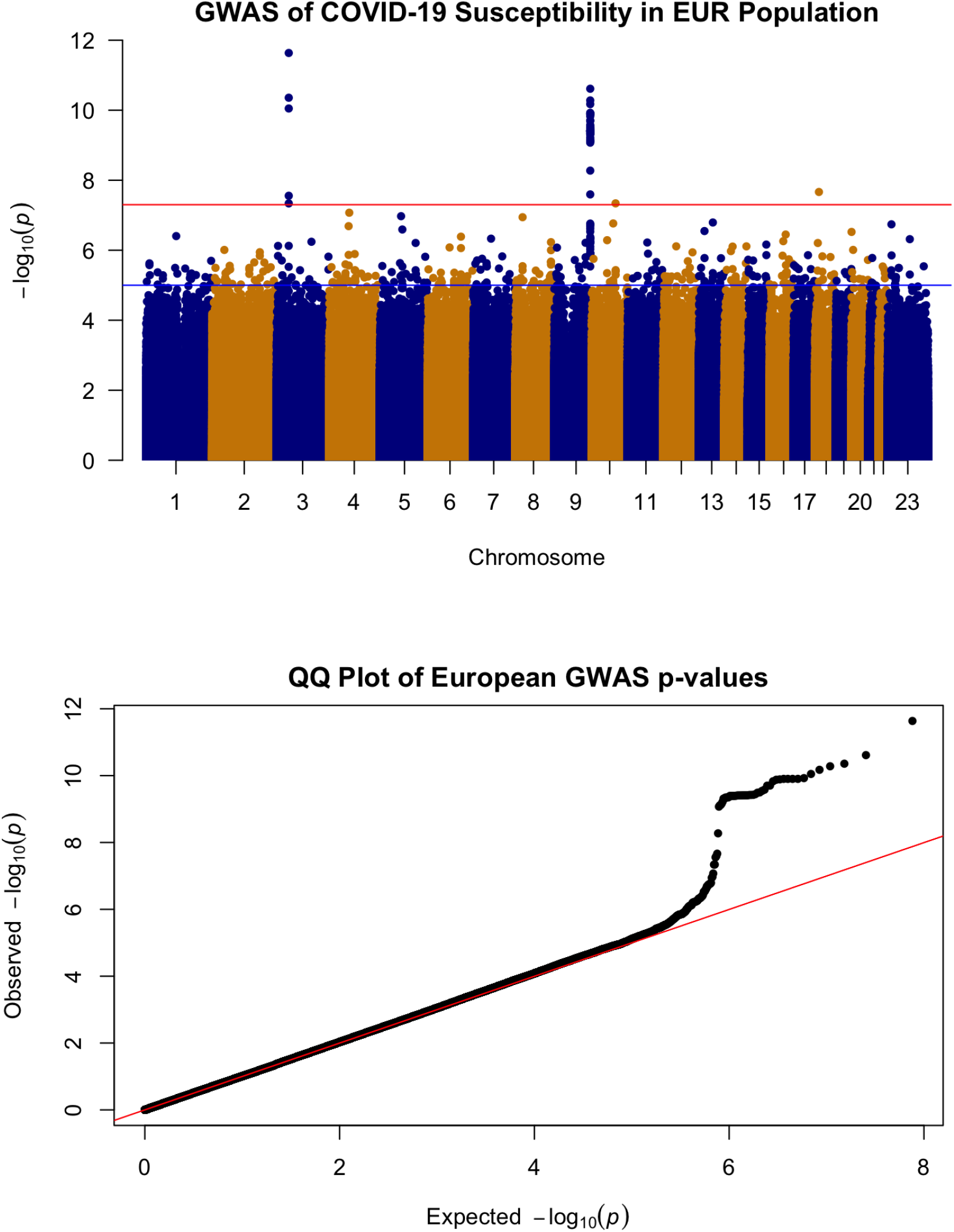
Manhattan and QQ plots for the analyzed associations in the UKB European population for COVID-19 susceptibility. The Manhattan plot’s red horizontal line corresponds to a log-transformed genome-wide significance level of 5 × 10^-8, and the blue line represents a suggested level of 1 × 10^-5.

**Figure 2.**
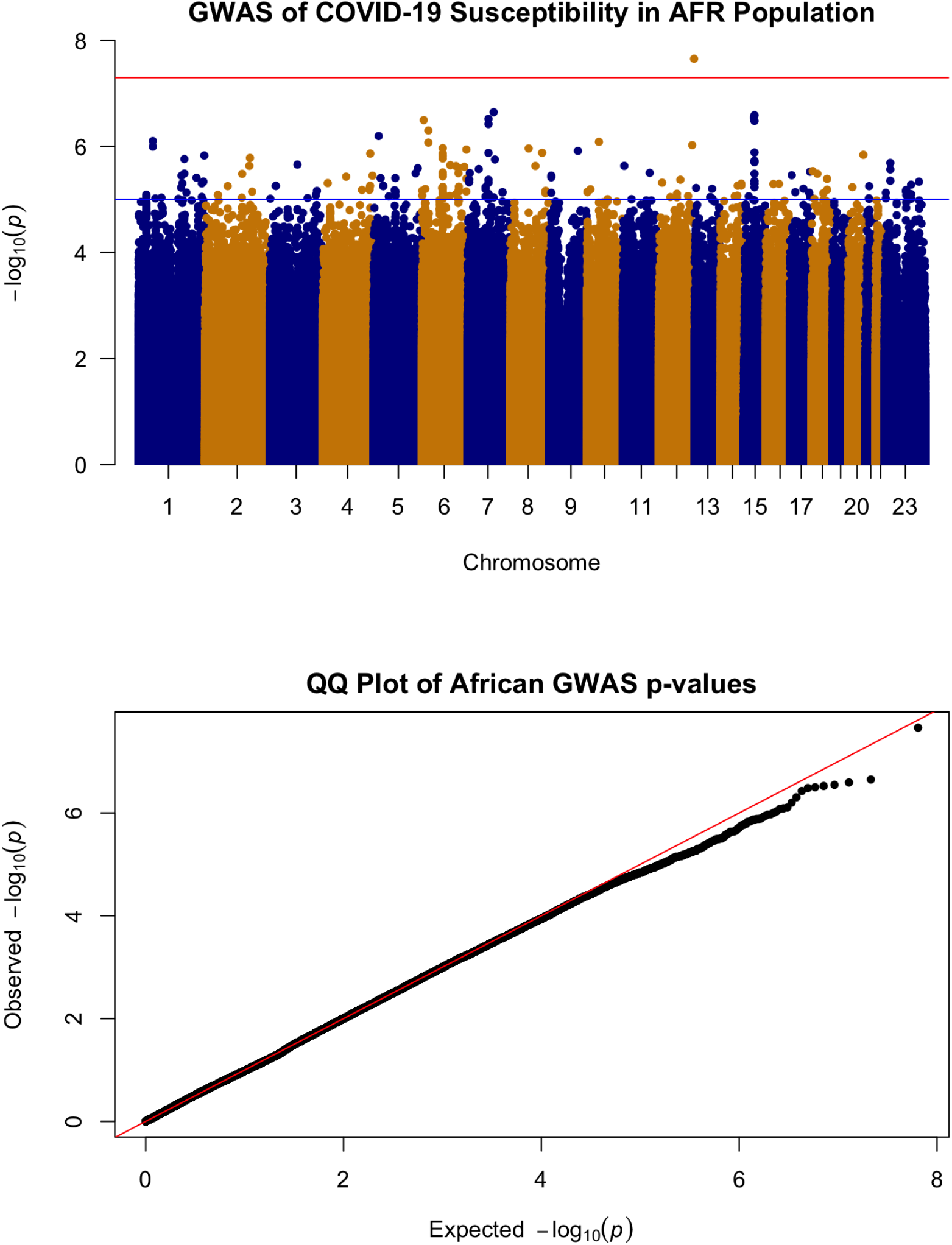
Manhattan and QQ plots for the analyzed associations in the UKB African population for COVID-19 susceptibility. The Manhattan plot’s red horizontal line corresponds to a log-transformed genome-wide significance level of 5 × 10^-8, and the blue line represents a suggested level of 1 × 10^-5.

**Figure 3.**
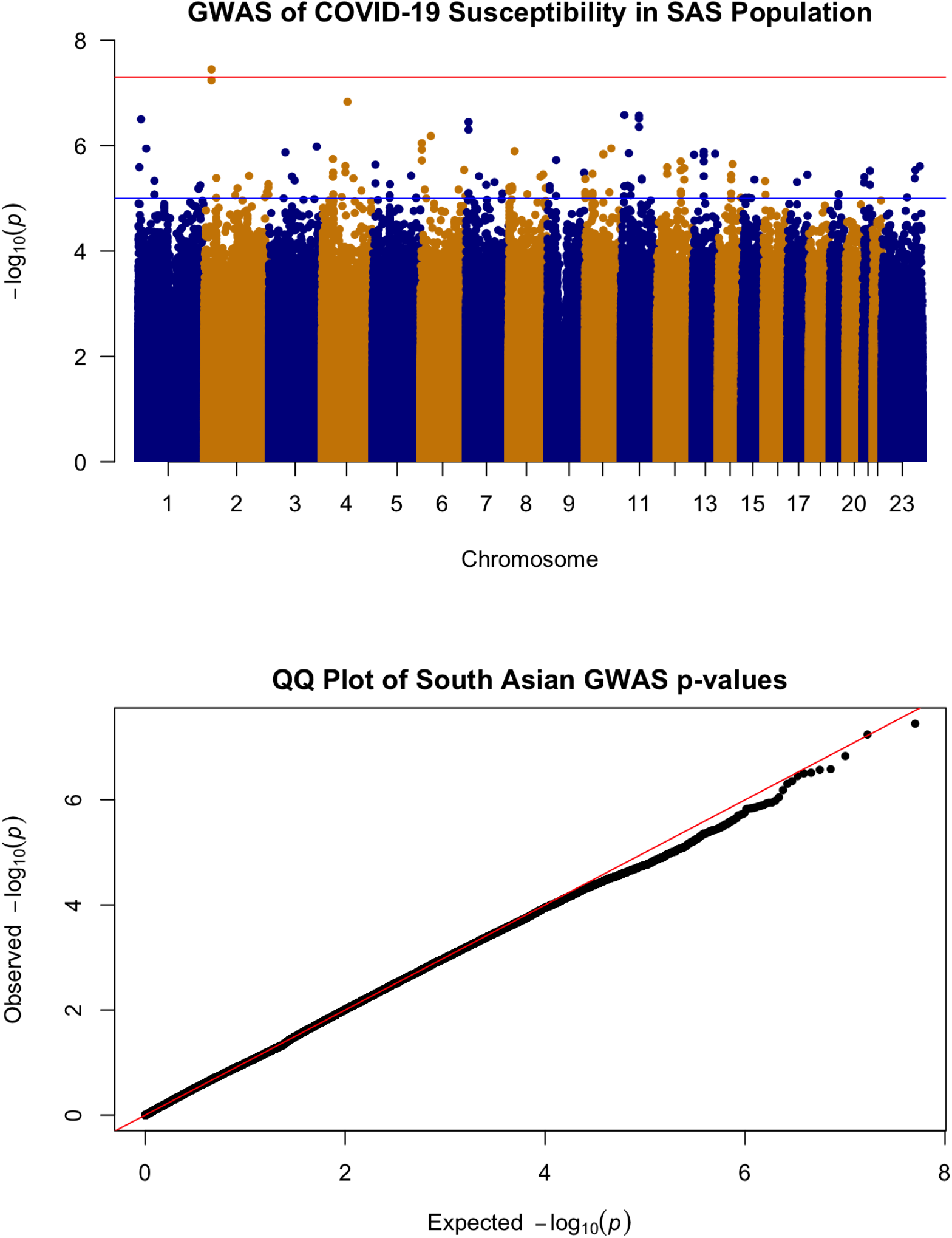
Manhattan and QQ plots for the analyzed associations in the UKB South Asian population for COVID-19 susceptibility. The Manhattan plot’s red horizontal line corresponds to a log-transformed genome-wide significance level of 5 × 10^-8, and the blue line indicates a suggested level of 1 × 10^-5.

**Figure 4.**
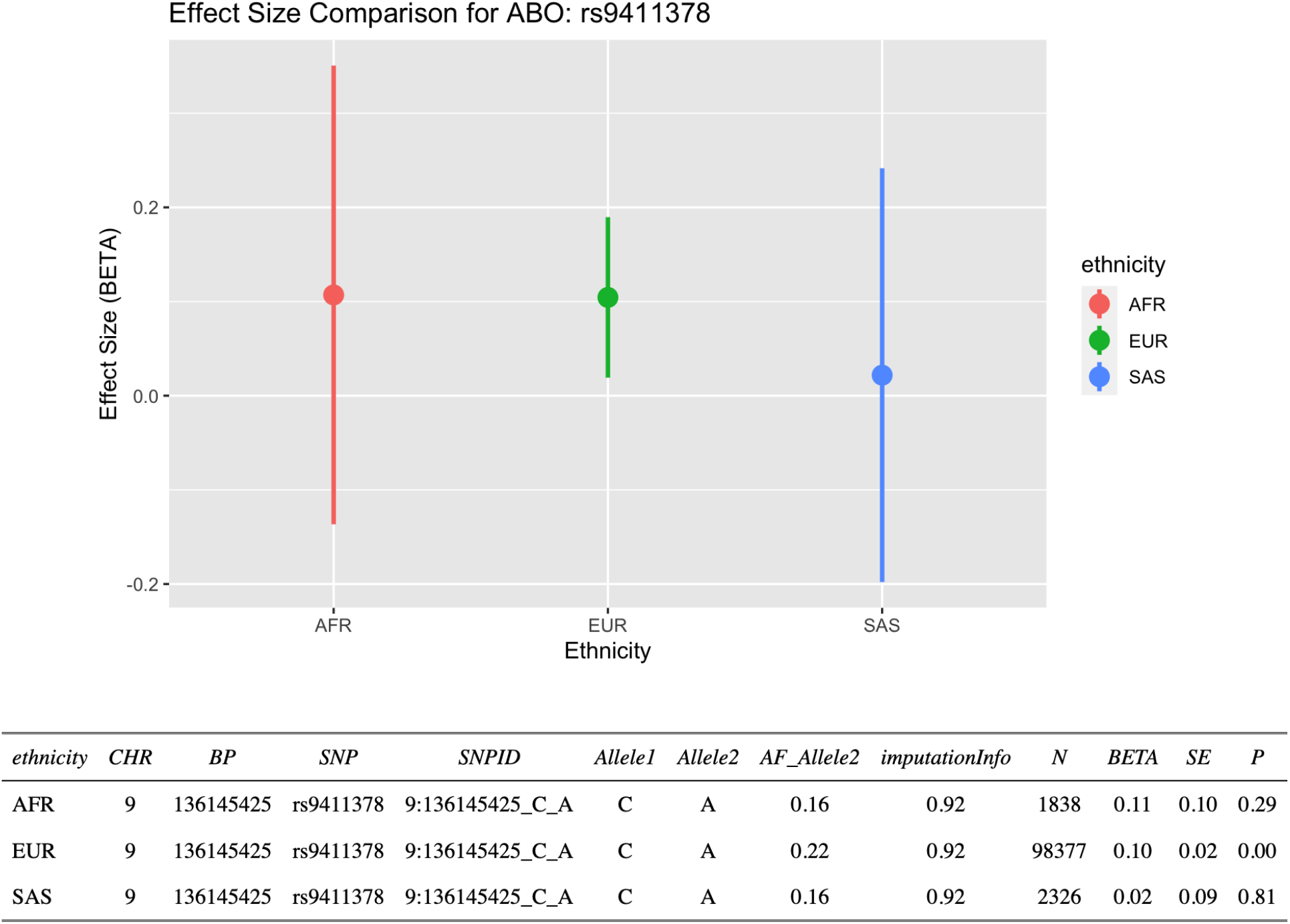
Comparison of associations (regression estimated β values) for rs9411378 with SARS-CoV-2 susceptibility. Vertical bars represent confidence intervals using α-levels that incorporate appropriate adjustments for multiplicity. For Europeans, α = 5×10-8 and for African ancestry and South Asians α = 0.05/3. The table displays selected summary statistics for the association between rs9411378 and SARS-CoV-2 susceptibility in all three ancestral populations of interest. *AF_Allele2* is the allele frequency of Allele2.

**Figure 5.**
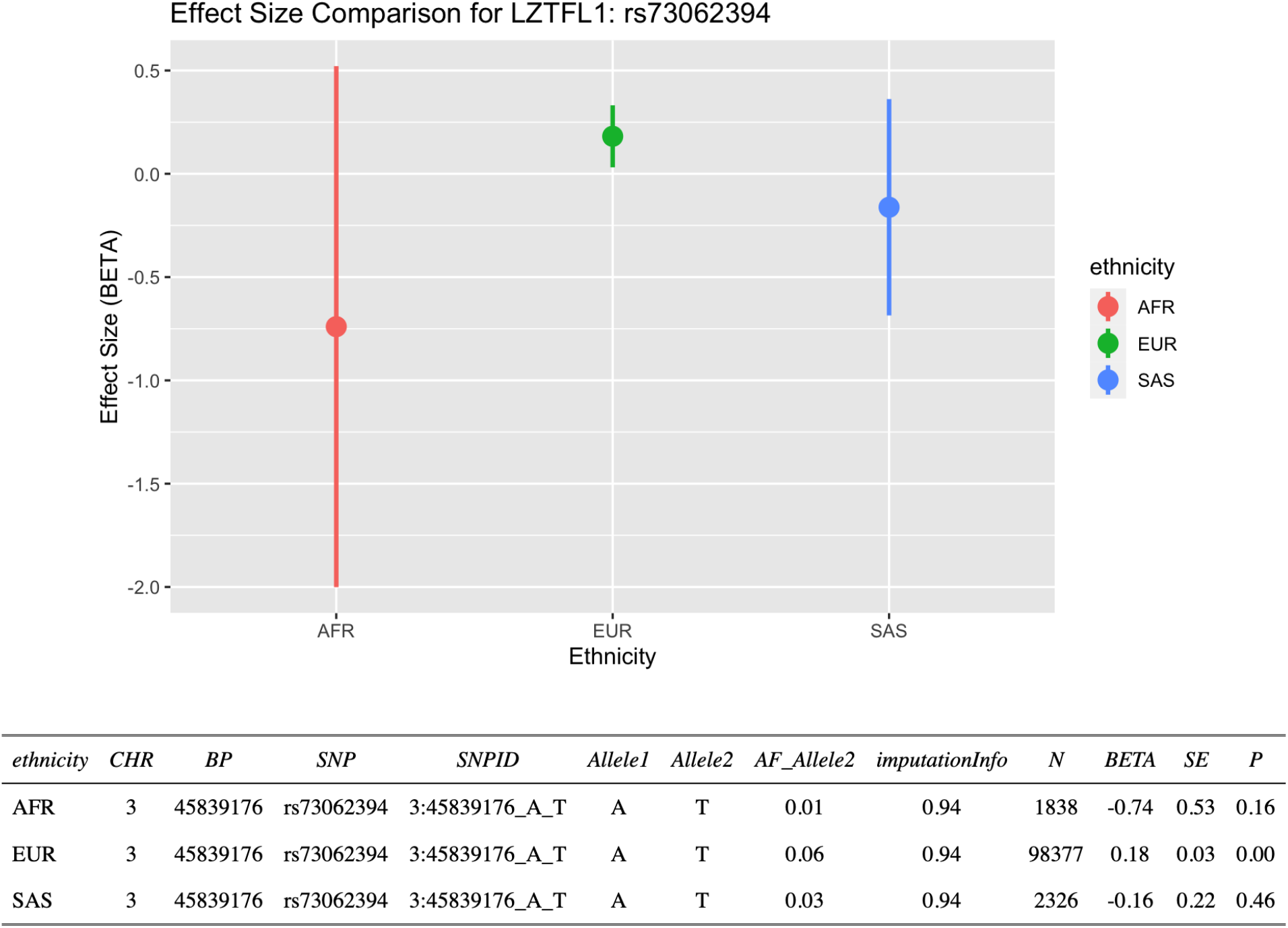
Comparison of associations (regression estimated β values) for rs73062394 with SARS-CoV-2 susceptibility. Vertical bars represent confidence intervals using α-levels that incorporate appropriate adjustments for multiplicity. For Europeans, α = 5×10-8 and for African ancestry and South Asians α = 0.05/3. The table displays selected summary statistics for the association between rs73062394 and SARS-CoV-2 susceptibility in all three ancestral populations of interest. *AF_Allele2* is the allele frequency of Allele2.

**Figure 6.**
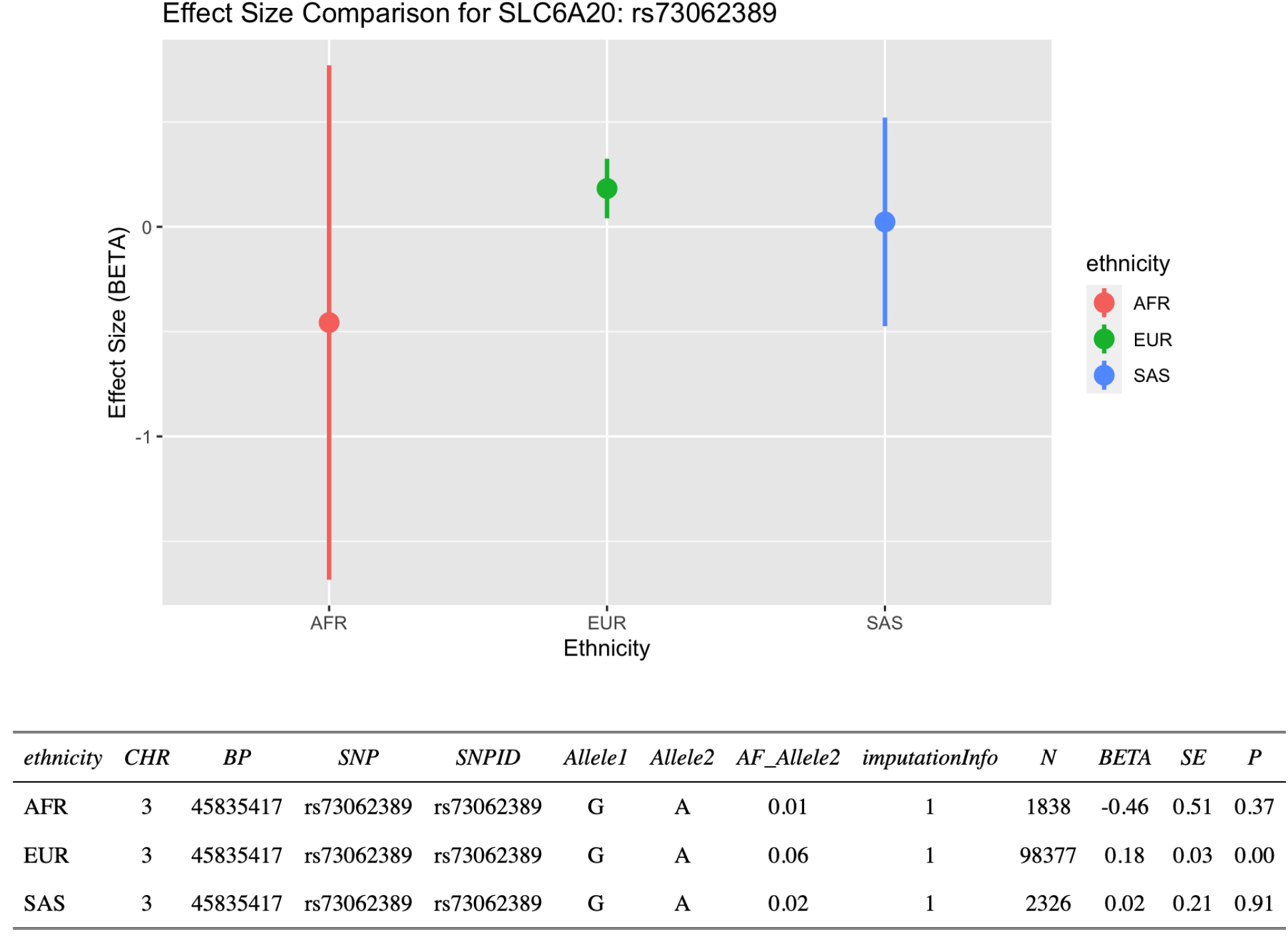
Comparison of associations (regression estimated β values) for rs73062389 with SARS-CoV-2 susceptibility. Vertical bars represent confidence intervals using α-levels that incorporate appropriate adjustments for multiplicity. For Europeans, α = 5×10-8 and for African ancestry and South Asians α = 0.05/3. The table displays selected summary statistics for the association between rs73062389 and SARS-CoV-2 susceptibility in all three ancestral populations of interest. *AF_Allele2* is the allele frequency of Allele2.

I assessed whether SNP associations with SARS-CoV-2 positive test results from the GWAS in European ancestry generalized to African and South Asian ancestry groups. For a given locus identified in the European ancestry GWAS, I selected the SNP with the smallest p-value to examine for replication. Additionally, for SNPs identified in the European ancestry GWAS, I compared the size and directionality of effect sizes (beta coefficient values) in the other two ancestry groups. Confidence levels for the regression estimated beta values were adjusted by their α-levels accordingly to account for multiplicity. For the European population, α = 5 × 10^−8^ for all confidence intervals for each SNP. For South Asian and African ancestries, α = 0.05/m for each SNP; in this Bonferroni adjustment to account for multiple comparisons, m represents the number of SNPs carried forward for replication.

## Results

Three loci in the European group had associated SNPs that were genome-wide significant (P < 5 × 10^−8^): *SLC6A20* in the chr3p21.31 locus (rs73062389-A; P = 2.315 × 10^−12^), *ABO* on chromosome 9 (rs9411378-A; P = 2.436 × 10^−11^) and *LZTFL1* on chromosome 3 (rs73062394; P = 4.401 × 10^−11^).^15, 16, 17^ These loci have been implicated in prior COVID-19 GWAS studies as potential modulators for SARS-CoV-2 infection susceptibility.^18, 19, 20^ A large phenome-wide association study for COVID-19 showed that the chr3p21.31 locus had a strong association with SARS-CoV-2 susceptibility due to its role in compromised lung tissue function.^6^ Additionally, data from a whole-lung RNA sequencing analysis revealed that *LZTFL1* is differentially over-expressed in the lung.^18^ The biological context for the discovered loci merits further investigation into the linkage disequilibrium and inheritance patterns of these SNPs.

I selected the three SNPs found in the EUR group for replication in the SAS and AFR populations; in both populations, these signals were not found to be genome-wide significant, possibly due to statistical power concerning the small sample sizes. Manhattan plots for each ancestral population are shown in Figures 1, 2, and 3 in tandem with the QQ plots.

Beta effects were further investigated as a metric of comparison since sample sizes differed considerably across the three ethnic groups of interest. Beta coefficient values represent log-odds ratios. For rs9411378 corresponding to the *ABO* locus, both the AFR and EUR populations had similar effect sizes and positive directionality (AFR beta coefficient = 0.107, SE = 0.102; EUR beta coefficient = 0.104, SE = 0.016). For rs73062394 corresponding to the *LZTFL1* locus, both the AFR and SAS populations had effect sizes with negative directionality (AFR beta coefficient = −0.740, SE = 0.526; SAS beta coefficient = −0.162, SE = 0.219). These directions of the effect estimates were opposite to that of the effect estimate for the EUR ancestry group (beta coefficient = 0.181, SE = 0.027). For rs73062389 corresponding to the *SLC6A20* locus, both the EUR and SAS populations had similar effect size and positive directionality (EUR beta coefficient = 0.183, SE = 0.031; SAS beta coefficient = 0.022, SE = 0.202).

## Discussion

With greater emphasis on increasing study participation in non-European populations, there will be increased opportunities to further genetic epidemiology methodology regarding how disease and genetics play connected roles across a wider variety of populations. Epidemiology will continue to play a vital role in our understanding of pandemics and outbreaks, and with the help of increased diversity in GWAS, scientists and clinicians will be able to inform more people on the proper courses of action to take to mitigate genetically-driven health consequences. Knowing, for example, that certain risk alleles may influence an increased likelihood of infection can impact vaccination and behavioral decisions. COVID-19 research, concerning a global pandemic affecting individuals and families everywhere, can benefit greatly from improved GWAS diversity.

For this investigation, I used three ancestral populations to examine associations between genetic variants and SARS-CoV-2 susceptibility. I discovered associations between SNPs at the *ABO, LZTFL1*, and *SLC6A20* loci in the European cohort that have been elucidated in prior GWAS papers regarding COVID-19. Despite smaller sample numbers in the African and South Asian populations, there were both effect size value and positive directionality similarities for rs9411378 at the *ABO* locus in the European and African groups. Additionally, there was a negative effect size directionality for rs73062394 at the *LZTFL1* locus in the African and South Asian groups. These findings prompt further investigation into SARS-CoV-2 and the genetic determinants driving variability in infection and infection response.

The alpha 1-3-N-acetylgalactosaminyltransferase (ABO) gene encodes the protein responsible for the human blood type determining system.^17^ Prior GWAS examining the association between rs9411378, the lead variant corresponding to the ABO locus, and COVID-19 susceptibility determined that the association remained even after controlling for other diseases including cardiovascular conditions and asthma. This suggested that confounding was not extensively involved in the association between the ABO locus and COVID-19 susceptibility.^5^ Additional GWAS for the severity phenotype also identified associations near ABO.^23^ The leucine zipper transcription factor like 1 (LZTFL1) gene encodes a cytoplasm-localized protein and helps regulate protein trafficking to ciliary membranes by interacting with Bardet-Biedl Syndrome proteins.^16^ A previous RNA-sequencing study identified increased *LZTFL1* expression in ciliated epithelial cells, and these cells are primary cellular targets during SARS-CoV-2 infection.^18, 22, 24^ Findings from our GWAS, however, showed negative effect sizes for the African and South Asian populations for rs73062394 at the *LZTFL1* locus; this may hint at a protective role for this locus in these ancestral groups. More evidence with larger population sample sizes is required to better understand *LZTFL1* and its role in SARS-CoV-2 infection. The solute carrier family 6 member 20 (SLC6A20) gene encodes a protein that functions as a proline transporter.^15^ This protein has been found to interact functionally with the ACE2 locus, which has been identified as a SARS-CoV-2 receptor.^21^

Findings from this GWAS as well as previous SARS-CoV-2 association studies are important, but must be interpreted with caution. Including diverse ancestries in a GWAS like this brings up study limitations with statistical power, due to the smaller sample sizes in the non-European ancestral groups. Furthermore, there has not been extensive clinical reporting of the SNPs found in this GWAS due to the recent nature of SARS-CoV-2 infection and the vast heterogeneity of clinical phenotypes resulting from infection. Future studies are required to further understand the genetic and overarching biological implications of these findings.

## Data Availability

All data produced in the present study are available upon reasonable request; contact

https://grasp.nhlbi.nih.gov/Covid19GWASResults.aspx

